# Left ventricular diastolic dysfunction attenuates outcomes in chronic thromboembolism pulmonary hypertension

**DOI:** 10.1101/2025.10.03.25334575

**Authors:** Katherine L Bunclark, Aleksandra Bartnik, Andrew Fletcher, Rob J van der Geest, Michael Newnham, John E Cannon, Gerry Coghlan, James Lordan, Luke Howard, David Jenkins, Martin Johnson, David G Kiely, Choo Ng, Karen Sheares, Dolores Taboada, Steven Tsui, S John Wort, Jonathan Weir-McCall, Joanna Pepke-Zaba, Mark Toshner

**Affiliations:** PVDU, Royal Papworth Hospital, Cambridge, U.K.; Leiden University Medical Center, Leiden, Netherlands; Heartlands Hospital, Birmingham, UK; Pulmonary Hypertension service, Royal Free Hospital, London, UK; Pulmonary Vascular Unit, Freeman Hospital, Newcastle Upon Tyne, UK; Pulmonary Hypertension service, Hammersmith Hospital, London, UK; Scottish Pulmonary Vascular Unit, Golden Jubilee National Hospital, Glasgow, UK; Sheffield PVDU, Royal Hallamshire Hospital, Sheffield, UK; Pulmonary Hypertension Service, Royal Brompton Hospital, London, UK; VPD Heart & Lung Institute, University of Cambridge, UK

## Abstract

**Background:** Left ventricular diastolic dysfunction in chronic thromboembolic pulmonary hypertension is classically attributed to the negative effects of pulmonary hypertension on left ventricular filling. Recent evidence, however, suggests diastolic dysfunction may exist independent of pulmonary hypertension and moreover may be masked by an under-filled left ventricle.

**Methods:** Consecutive patients undergoing pulmonary endarterectomy (2007 – 2018) were included (n=1266). Left ventricular diastolic dysfunction was assessed using pulmonary arterial wedge pressure and in nested cohorts utilising multi-modal cardiac imaging. Diagnostic baseline, outcome data, and long-term mortality outcomes were assessed.

**Results:** 135 individuals had a wedge pressure >15mmHg following surgery, of whom 60% had a normal wedge pressure pre-operatively. No patients had a formal diagnosis of heart-failure preserved ejection fraction. Haemodynamic, functional and patient-related outcomes were all worse in this patient subgroup and associated with a higher requirement for peri-operative non-invasive ventilation and impaired long-term survival. Post-operative cardiac imaging confirmed evidence of left ventricular diastolic dysfunction in patients with an elevated wedge pressure. Pre-operative left atrial dilatation alone predicted post-operative wedge elevation with accuracy (sensitivity 67%, specificity 100%), and was superior to echocardiography.

**Conclusions:** Left ventricular diastolic dysfunction is strongly associated with all pulmonary endarterectomy outcome measures, however the majority of patients do not have an elevated wedge pressure pre-operatively and are not diagnosed with heart failure. Standard pre-operative work-up should include assessment for diastolic dysfunction to aid risk categorisation and guide therapy decisions.

## Introduction

Chronic thromboembolic pulmonary hypertension (CTEPH) is an infrequent but important complication of pulmonary embolism. Organisation and fibrotic transformation of thromboembolic material in the pulmonary arteries, together with a secondary small-vessel vasculopathy, gives rise to pulmonary hypertension (PH) [1]. Pulmonary Endarterectomy (PEA) is the guideline recommended treatment in those with surgically accessible disease and results in symptomatic improvement and survival [2–4]

Left untreated, CTEPH results in progressive right ventricular (RV) overload with morphological and functional abnormalities affecting both ventricles [5]. Whilst left ventricular (LV) systolic function in CTEPH is normally preserved, LV diastolic function is frequently impaired [6–9]. Abnormalities of mitral inflow in early diastole are most often observed, presumably due to the negative effects of PH on LV preload, geometry and isovolumetric relaxation [5–12].

Further support for the concept of this secondary LV diastolic dysfunction (LVDD) in CTEPH is lent by the rise in LV end-diastolic pressure (LVEDP) and normalisation of LV filling patterns following PEA [6, 12–14]. Despite this, ~ 11% of CTEPH patients have elevated LV filling pressures on invasive haemodynamic assessment, independent of measures of PH severity, and strongly associated with left atrial dilatation [14]. Primary LVDD may therefore be present in a proportion of those with CTEPH, irrespective of the contributory effect of PH on LV filling dynamics and moreover may be masked by an under-filled LV [14, 15].

Elevated LVEDP, by invasively-derived and cardiac MR measures, is a recognised independent risk factor for long-term survival in CTEPH [14, 16]. Its effect on mortality and morbidity outcomes following PEA are unknown. Furthermore, the ability to identify individuals with primary LVDD from pre-operative measures has yet to be explored. With this in mind, we provide the largest multi-centre evaluation of LVDD in operable CTEPH and its effect on patient outcomes post-PEA.

## Materials and methods

### Patient Selection

Consecutive individuals undergoing PEA for CTEPH at the National PEA Centre (Royal Papworth Hospital, United Kingdom) from August 2007 - December 2018 were included. Patient selection is summarised in Figure 1. CTEPH diagnosis was as per international criteria at time of surgery, with PEA performed as previously described [4, 17]. The ethic committee of Royal Papworth Hospital, UK waived ethical approval for this study. Patient data was de-identified prior to use in this study.

**Figure 1:**
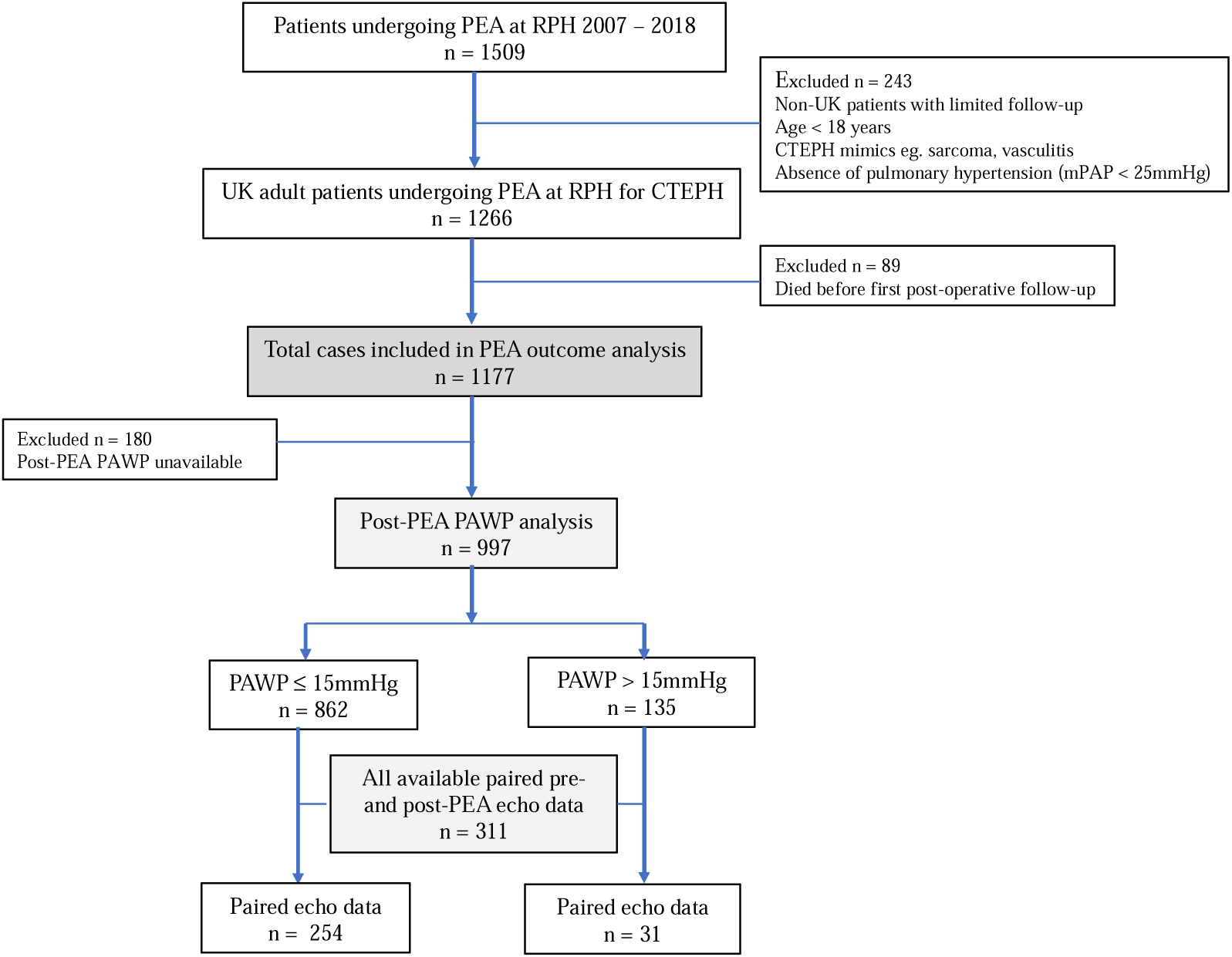
Study consort diagram including inclusion/exclusion criteria. PEA, Pulmonary Endarterectomy; UK, United Kingdom; RPH, Royal Papworth Hospital; CTEPH, Chronic thromboembolic pulmonary hypertension; mPAP, mean pulmonary artery pressure; PAWP, pulmonary arterial wedge pressure

### Patient Characteristics and Outcomes

Pre- and post-PEA data were obtained from a prospectively curated local database. Patient-reported outcomes (PROs) were assessed using the Cambridge Pulmonary Hypertension Outcome Review (CAMPHOR) [18]. Pre-operative data was taken at diagnosis and outcome data at first post-operative follow-up 3 – 6 months following PEA. Non-invasive tests were contemporaneous to invasive haemodynamics. Survival was censored at 31^st^ March 2022 from a centralised national resource.

### Invasively-derived pulmonary artery wedge pressures

Pulmonary Artery Wedge Pressure (PAWP) was dichotomised into ≤ 15mmHg and > 15mmHg groups based upon current thresholds in CTEPH [17]. Right heart catheterisation and PAWP derivation was undertaken by an experienced PH Physician as in [19, 20]. PAWP was utilised over LVEDP due to the requirement for additional invasive procedures above standard PEA clinical care in attaining LVEDP.

### Echocardiography

Transthoracic echocardiograms were analysed for a subset of the cohort with paired pre- and post-operative echocardiographic data, contemporaneous to invasive haemodynamics.. Recorded echocardiographic parameters are detailed in Table S1. PH and LVDD were assessed as per guidelines [21, 22]. Left atrial size was recorded as area rather than volume in line with reporting standards at the time. There were no instances of severe valvular disease.

### Cardiac Magnetic Resonance Imaging (CMR)

CMR images were analysed for a subset of the cohort with paired pre- and post-operative CMR data available. CMR images were obtained at the referring centres using 1.5-T cardiac optimised system with post-processing performed as described in [23].

### Statistical Analysis

Statistical analysis was performed using R version 3.6.1 [24]. Data averages for continuous variables were reported as median +/− interquartile range (IQR) and categorical variables as n (percentage of total). Groups were compared using Mann Whitney *U* test for continuous data and Chi-squared test with Yates correction for categorical data. Reported p-values were adjusted for multiple comparisons by False Discovery Rate at 5%. Multivariable Cox proportional hazards models were used to calculate hazards ratios and 95% confidence intervals for long-term survival. Additional methodology is provided in the online supplement.

## Results

### Cohort patient characteristics

1266 individuals with CTEPH underwent PEA during the study period. Cohort characteristics are summarised in Table 1. Median ± IQR age at PEA was 62 ± 22 years and 54% were male. PEA resulted in significant improvements in all haemodynamic, functional and patient-reported outcome (PRO) measures (p ≤ 0.001, all; Table S2). Median time from PEA to outcome assessment was 138 ± 82 days.

**Table 1:** Patient characteristics and demographics at pre-PEA baseline.

|  | n | All PEA<br>(n=1266) | n | PAWP<br>≤15mmHg<sup>a</sup><br>(n=862) | n | PAWP<br>> 15mmHg<sup>a</sup><br>(n=135) | Adj p-value |
| --- | --- | --- | --- | --- | --- | --- | --- |
| **Age at PEA, yrs** | 1266 | 62 ± 22 | 862 | 61 ± 21 | 135 | 63 ± 21 | 0.422 |
| **Male sex, n (%)** | 1266 | 681 (54) | 862 | 476 (55) | 135 | 73 (54) | 0.991 |
| **BMI, kg/m<sup>2</sup>** | 909 | 29 ± 8 | 693 | 28 ± 7.0 | 107 | 33 ± 8.5 | < 0.001 |
| **FEV1/FVC, ratio** | 533 | 72 ± 12 | 378 | 0.73 ± 0.1 | 45 | 0.71 ± 0.1 | 0.478 |
| **Smoker,<sup>b</sup> n (%)** | 912 | 471 (52) | 683 | 348 (51) | 113 | 65 (58) | 0.441 |
| **Comorbidities, n (%)** |  |  |  |  |  |  |  |
| Atrial arrhythmia | 1227 | 115 (9) | 839 | 66 (7.8) | 132 | 24 (18) | < 0.001 |
| Systemic HTN | 1227 | 334 (27) | 839 | 225 (27) | 132 | 48 (36) | 0.107 |
| Type 2 Diabetes | 1227 | 135 (11) | 838 | 76 (9) | 132 | 23 (17) | 0.023 |
| IHD <sup>c</sup> | 1245 | 155 (12) | 854 | 92 (11) | 133 | 23 (17) | 0.133 |
| **Haemodynamics** |  |  |  |  |  |  |  |
| Mean PAP, mmHg | 1266 | 45 ± 15 | 862 | 44 ± 16 | 135 | 46 ± 14 | 0.464 |
| PVR, dynes | 1202 | 676 ± 485 | 862 | 669 ± 495 | 135 | 555 ± 403 | 0.005 |
| PAWP, mmHg | 1052 | 11 ± 5 | 862 | 11 ± 6 | 135 | 14 ± 6 | < 0.001 |
| CI, l/min/m2 | 1125 | 2.1 ± 0.75 | 862 | 2.1 ± 0.76 | 135 | 2.2 ± 0.75 | 0.711 |
| **Functional Status** |  |  |  |  |  |  |  |
| NYHA, 1/2/3/4% | 1088 | 0/15/74/11 | 796 | 0/16/73/10 | 125 | 0/10/77/14 | 0.204 |
| 6MWD, metres | 732 | 316 ± 195 | 555 | 320 ± 183 | 67 | 240 ± 218 | 0.002 |
| **Intra-operative** |  |  |  |  |  |  |  |
| CPB time, mins | 1266 | 321 ± 65 | 839 | 320 ± 65 | 127 | 324 ± 60 | 0.353 |
| DHCA time, mins | 1266 | 37 ± 16 | 783 | 37 ± 15 | 113 | 37 ± 16 | 0.547 |
| **Other surgery, n (%)** |  |  |  |  |  |  |  |
| All additional | 1266 | 159 (13) | 862 | 97 (11) | 135 | 24 (18) | 0.082 |
| CABG | 1266 | 95 (8) | 862 | 56 (6.5) | 135 | 15 (11) | 0.230 |
| AVR | 1266 | 14 (1) | 862 | 7 (0.8) | 135 | 3 (2) | 0.478 |
| MVR | 1266 | 9 (0.7) | 862 | 5 (0.6) | 135 | 2 (2) | 0.722 |
| ASD/PFO closure | 1266 | 5 (0.4) | 862 | 32 (4) | 135 | 4 (3) | 1.00 |
| **Complications, n (%)** |  |  |  |  |  |  |  |
| CPAP | 1136 | 292 (26) | 767 | 168 (22) | 128 | 44 (34) | 0.015 |
| Haemofiltration | 1075 | 62 (6) | 719 | 24 (3) | 119 | 8 (7) | 0.342 |
| ECMO | 1201 | 64 (5) | 815 | 18 (2) | 126 | 2 (2) | 0.991 |
| Pneumonia | 1131 | 167 (15) | 766 | 90 (12) | 126 | 22 (18) | 1.00 |
| Return to theatre | 1135 | 82 (7) | 770 | 45 (5) | 122 | 10 (8) | 0.644 |
| Reperfusion injury | 1136 | 86 (8) | 767 | 49 (6) | 125 | 7 (6) | 0.991 |
| **Intubation, days** | 1173 | 1 ± 1 | 809 | 1 ± 1 | 131 | 1 ± 1 | 0.844 |
| **Hospital stay, days** |  |  |  |  |  |  |  |
| Intensive care unit | 1237 | 3 ± 4 | 840 | 3 ± 3 | 133 | 3 ± 4 | 0.422 |
| Total inpatient | 1237 | 12 ± 9 | 839 | 12 ± 8 | 132 | 13 ± 11 | 0.005 |

Data are median ± IQR unless stated otherwise. Pre-operative values taken at closest time-point to diagnostic right heart catheterization. Percentages may not add to 100 due to rounding. Reported p-values are adjusted for multiple comparisons with false discovery rate. PEA; Pulmonary Endarterectomy; PAWP: Pulmonary Arterial Wedge Pressure; BMI: Body Mass Index; FEV1: Forced Expiratory Volume in one second; FVC: Forced Vital Capacity; HTN: hypertension; IHD: Ischaemic Heart Disease; PAP: Pulmonary Artery Pressure; PVR: Pulmonary Vascular Resistance; PAWP: Pulmonary Arterial Wedge Pressure; CI: Cardiac Index; NYHA: New York Heart Association class; 6MWD: Six-minute walk distance; CPB: Cardiopulmonary Bypass; DHCA: Deep Hypothermic Circulatory Arrest; CABG : Coronary Artery Bypass Grafting; AVR: Aortic Valve Replacement; MVR; Mitral Valve Replacement; ASD: Atrial Septal Defect; PFO: Patent Foramen Ovale; CPAP: Continuous Positive Airways pressure; ECMO: Extracorporeal Membrane Oxygenation.
<sup>a</sup> PAWP taken at first follow-up within one year of PEA
<sup>b</sup> Current or previous tobacco smoker
<sup>c</sup> History of angina, coronary artery stenting, use of nitrates or coronary artery bypass grafting required at time of pulmonary endarterectomy

### Echocardiography in CTEPH

Patient characteristics in the echocardiography sub-cohort (n = 311) were similar to the main cohort (Table S3). Prior to PEA, LV ejection fraction was preserved (64 ± 5%). LV filling was abnormal with reduced E-wave velocity (55.6 ± 25.1 cm/s) and E/A reversal (0.75 ± 0.29). LV relaxation by lateral é and E/é, left atrial area and LV mass index were within normal limits (Table S1). There were significant improvements in LV filling and compliance measures (left atrial area, E-wave velocity, E/A and E/é ratios, tricuspid regurgitation velocity and mitral deceleration time (p < 0.001, all) with PEA. Lateral é was unchanged (p = 0.457).

### CMR in CTEPH

The CMR sub-cohort comprised of 142 individuals (n = 90 with contemporaneous paired echocardiography). Patient characteristics were similar to the main cohort (Table E4). With sex differences accounted for, LV end diastolic/systolic volumes, mass, ejection fraction and cardiac output were within normal limits prior to PEA whilst LV stroke volume, left atrial volume and left atrial ejection fraction were reduced and LV mass/volume increased (Table S5). PEA resulted in a rise in LV stroke volume (p = 0.032), LV end-diastolic (p < 0.001) and end-systolic (p = 0.032) volumes and cardiac output (p < 0.001). LV mass also increased (p = 0.017) whilst LV mass/volume decreased (p = 0.009). LV ejection fraction remained within normal limits (55 ± 10%; p = 0.041). Left atrial volume (p < 0.001) also rose whilst left atrial ejection fraction fell (p = 0.002). All measures of RV morphology and function improved following surgery (p < 0.05; Table S5).

### LV diastolic function post-PEA

Post-operative PAWP measurements were recorded for 997 individuals of whom 862 (86%) had a PAWP ≤ 15mmHg and 135 (14%) a PAWP > 15mmHg. There were 89 deaths prior to first post-operative RHC and PAWP assessment. Paired pre- and post-PEA PAWP readings were available for 836 individuals (Figure S1). 78% (n = 653) had a PAWP ≤ 15mmHg before and after surgery. 40% with an elevated PAWP post-PEA had a PAWP >15mmHg prior.

Cardiovascular co-morbidities including Type 2 Diabetes (p = 0.023) and obesity (p < 0.001) were more common in those with an elevated PAWP following PEA, as were chronic atrial arrhythmias (p < 0.001; Table 1). Those with PAWP elevation post-PEA had a higher pre-operative PAWP (p < 0.001), lower PVR (p = 0.005) and lower walk distance (p = 0.002). Intra-operative factors did not differ between PAWP groups. There were no distinguishing features between those with a normal and elevated pre-operative PAWP and an elevated post-operative PAWP (Table S6)

In the echocardiographic sub-cohort, 31 individuals (10.8%) had an elevated PAWP post-op. PAWP elevation was associated with left atrial dilatation and measures of LVDD but preserved LV systolic function (Table 2). Excluding those with atrial arrhythmias did not alter findings (Table S7). Post-operative PAWP elevation could be discerned pre-operatively by higher: left atrial area (p < 0.001), average E/é (p = 0.036), E (p = 0.003) and A-wave (p = 0.004) velocities and LV mass index (p = 0.003). Left atrial area alone was the best predictor of post-operative PAWP elevation with a threshold value of 17.8 mls in the absence of atrial arrhythmia (specificity 63.2%, sensitivity 77.3%).

**Table 2:** Echocardiographic variables at first post-operative RHC stratified by PAWP.

|  | **PAWP ≤ 15mmHg**<br>**(n=254)** | **PAWP > 15mmHg**<br>**(n=31)** | **p-value** |
| --- | --- | --- | --- |
| Left atrial area, cm<sup>2</sup> | 18.3 ± 5.4 | 22.8 ± 3.3 | 0.007 |
| Right atrial area, cm<sup>2</sup> | 19.5 ± 7.7 | 24.0 ± 11.0 | 0.002 |
| TR velocity, m/s | 260 83 | 305 40 | 0.005 |
| LVEF, % | 60 ± 6 | 60 ± 8 | 0.977 |
| LV mass index, g/m<sup>2</sup> | 72.6 ± 24.0 | 82.2 ± 19.4 | 0.018 |
| LV RWT | 0.40 ± 0.11 | 0.47 ± 0.06 | 0.018 |
| Mitral E, cm/s | 63.8 ± 28.1 | 90.0 ± 26.5 | < 0.001 |
| Mitral A, cm/s | 71.0 ± 27.0 | 97.0 ± 27.0 | 0.018 |
| Mitral E/A ratio | 0.85 ± 0.5 | 0.85 ± 0.4 | 0.501 |
| Mitral lateral e', cm/s | 10.9 ± 4.7 | 10.3 ± 4.6 | 0.977 |
| Mitral lateral E/e' | 6.10 ± 3.2 | 7.8 ± 1.7 | 0.004 |
| Mitral average E/e' | 8.0 ± 3.1 | 9.4 ± 3.6 | 0.018 |
| Mitral DT, ms | 210 ± 78 | 170 ± 50 | 0.018 |

Values expressed as median ± IQR.
PAWP, pulmonary artery wedge pressure; TR, tricuspid regurgitation; LVEF, Left ventricular ejection fraction;
LV, left ventricle; RWT, relative wall thickness; DT, deceleration time.

In the CMR sub-cohort, 14 individuals (10.5%) has an elevated post-operative PAWP (Table 3). PAWP elevation was associated with significantly lower LV end-diastolic (p = 0.032) and end-systolic (p = 0.048) volumes, lower LV stroke volume (p = 0.048) and a higher LV mass/volume ratio (p = 0.032) when compared to their normal PAWP counterparts. Left atrial volume was higher in the raised PAWP group (p = 0.017), though did not reach significance when indexed for body surface area.

**Table 3:** CMR variables at first post-operative RHC stratified by PAWP.

|  | **PAWP ≤**<br>**15mmHg**<br>**(n= 119)** | **PAWP >**<br>**15mmHg**<br>**(n=14)** | **p-value** |
| --- | --- | --- | --- |
| **Right atrium** |  |  |  |
| Volume index, mls/m<sup>2</sup> | 38 ± 24 | 44 ± 25 | 0.628 |
| **Right ventricle** |  |  |  |
| End-diastolic volume index, mls/m<sup>2</sup> | 77 ± 22 | 86 ± 22 | 0.782 |
| End-systolic volume index, mls/m<sup>2</sup> | 40 ± 15 | 50 ± 25 | 0.224 |
| Ejection fraction, % | 47 ± 13 | 38 ± 9 | 0.044 |
| Cardiac output, l/min | 5.4 ± 1.9 | 6.0 ± 2.5 | 0.881 |
| Stroke volume index, mls/m<sup>2</sup> | 38 ± 14 | 35 ± 13 | 0.307 |
| Mass index, g/m<sup>2</sup> | 20 ± 5 | 21 ± 3 | 0.308 |
| **Left atrium** |  |  |  |
| Volume, mls | 63 ± 28 | 76 ± 23 | 0.017 |
| Volume index, mls/m<sup>2</sup> | 32 ± 18 | 36 ± 11 | 0.230 |
| Ejection fraction, % | 57 ± 10 | 45 ± 8 | 0.006 |
| **Left ventricle** |  |  |  |
| End-diastolic volume index, mls/m<sup>2</sup> | 76 ± 17 | 59 ± 9 | 0.032 |
| End-systolic volume index, mls/m<sup>2</sup> | 33 ± 13 | 29 ± 10 | 0.048 |
| Ejection fraction, % | 55 ± 10 | 56 ± 8 | 0.801 |
| Cardiac output, l/min | 5.9 ± 1.73 | 5.5 ± 2.12 | 0.446 |
| Stroke volume index, mls/m<sup>2</sup> | 41 ± 12 | 34 ± 6 | 0.048 |
| Mass index, g/m<sup>2</sup> | 55 ± 18 | 60 ± 12 | 0.881 |
| Mass/Volume, g/ml | 0.8 ± 0.19 | 0.9 ± 0.42 | 0.032 |

Values expressed as median ± IQR.
PAWP, pulmonary artery wedge pressure

Those with a normal post-operative PAWP saw significant reductions in RV dilatation and hypertrophy with PEA and a significant increase in RV cardiac output and ejection fraction. RV stroke volume remained unchanged (Table S8). LV volumes, stroke volume and cardiac output also increased with surgery whilst LV mass/volume ratio fell. Left atrial volume significantly rose whilst left atrial ejection fraction fell. In comparison, for those with a high post-operative PAWP there were no significant change with PEA across any of the measures of RV, LV and left atrial measures of morphology and function (Table S9). Pre-operative left atrial volume alone was the strongest predictor of post-operative PAWP elevation with a threshold value of 63.1mls in the absence of atrial arrythmia (specificity 66.7%, sensitivity 100%).

### PEA outcomes

PEA resulted in significant improvements in haemodynamics and PRO measures irrespective of post-operative PAWP, with the exception of cardiac index (p = 0.393) and plasma NTproBNP level (p = 0.993) which remained unchanged in those with elevated post-operative PAWP (Figure 2 and Table S10). Despite observed improvements with PEA, post-operative PAWP >15mmHg was associated with significantly lower post-operative gains (mean pulmonary artery pressure, cardiac index, NYHA class, 6-minute walk distance, PROs and NTproBNP level, p < 0.001 all; Figure 2 and Table S11). An increased requirement for non-invasive ventilation post PEA (p = 0.015) was also observed in those with elevated PAWP, though rates of reperfusion lung injury and duration of invasive ventilation did not differ (Table 1).

**Figure 2:**
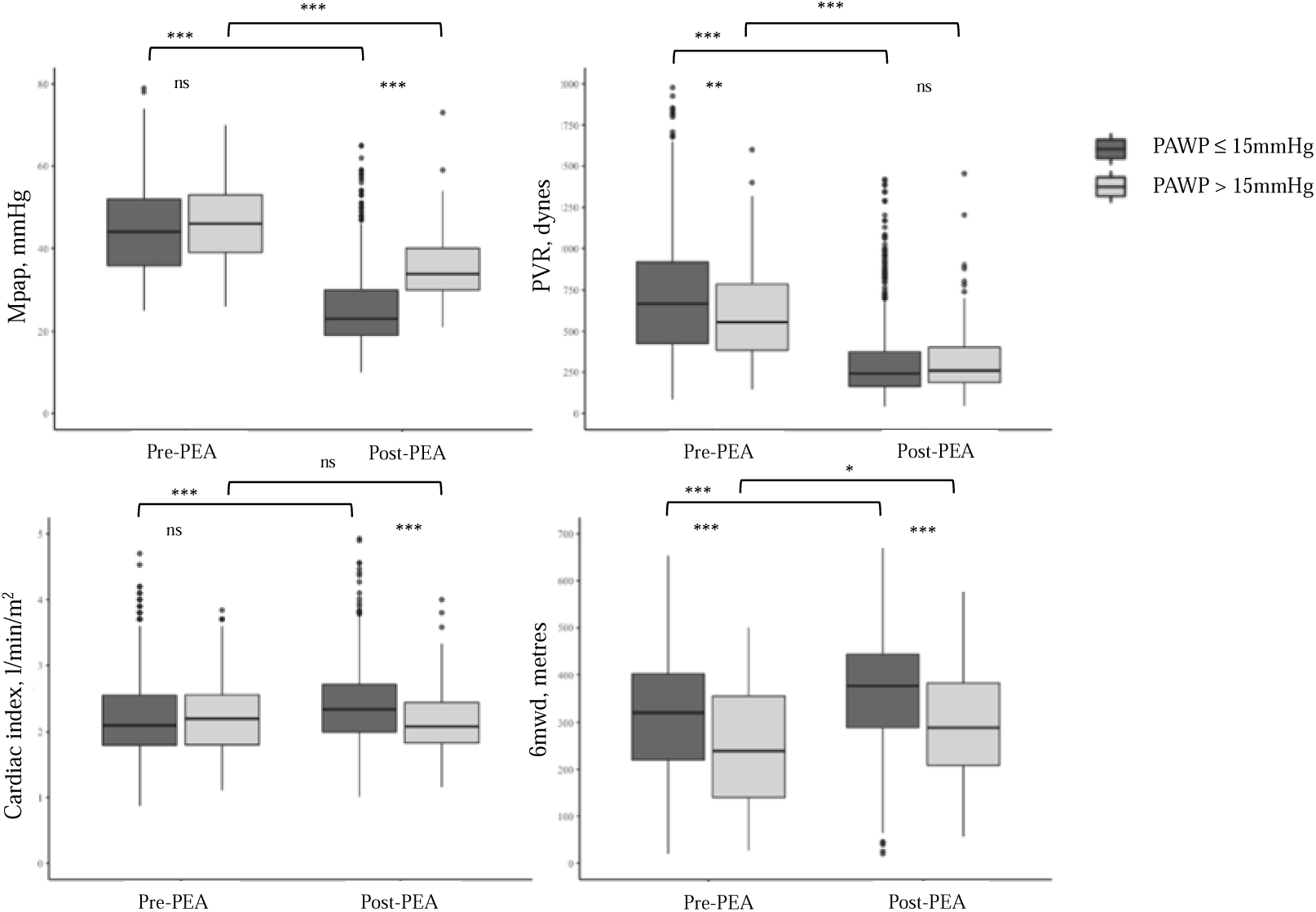
Key pulmonary endarterectomy outcome measures. *p ≤ 0.05; ** p ≤ 0.01; *** p ≤ 0.001; ns, not statistically significant P-values adjusted for multiple comparisons using false discovery rate controlled at 5%. MPAP, mean pulmonary artery pressure; PEA, pulmonary endarterectomy; PVR, pulmonary vascular resistance; 6MWD, six-minute walk distance.

### PEA survival

Conditional cohort survival at 1, 3 and five-years was 92.8%, 89.7%, 81.9% respectively. Individuals who died prior to first follow-up were older (p < 0.001), had worse PROs, high NYHA class and lower 6-minute walk distance (p < 0.001) prior to PEA (Table S12). No associations were found between pre-operative cardiac imaging measures and early death.

Individuals with an elevated PAWP had a 1, 3 and 5 year survival of 98.5%, 91.6% and 77.6% compared to 99.3%, 95.5% and 90.5% in those who did not (p < 0.001; Figure 3). Post-operative PAWP > 15mmHg was an independent predictor of mortality (Hazard Ratio [95% CI]), 2.01 [1.16 – 3.48]; p = 0.012) along with older age (1.04 [1.02 – 1.06]; p = 0.001), and lower 6-minute walk distance (0.41 [0.28 – 0.59]; p 0.029; Table S13).

**Figure 3:**
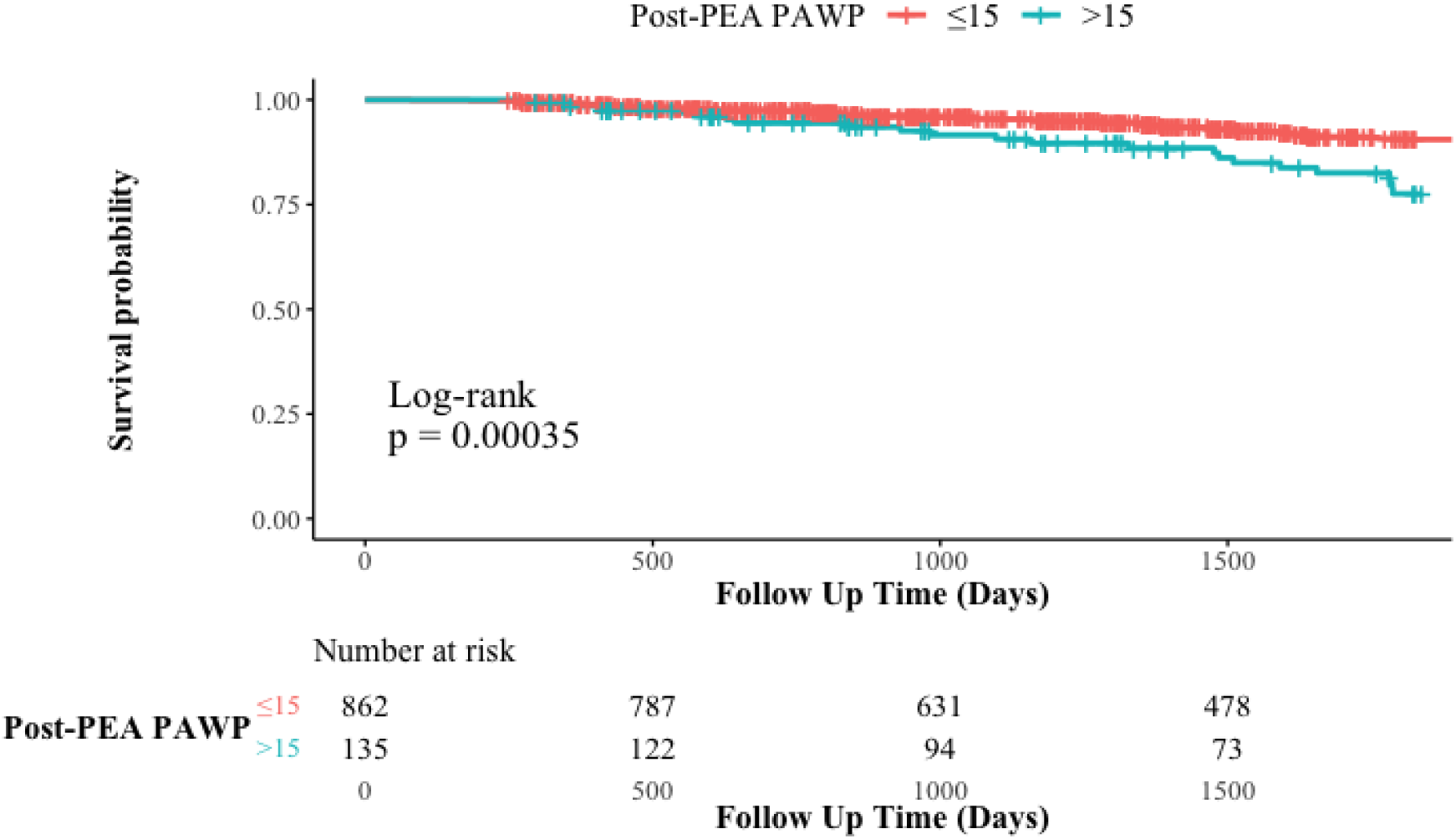
Kaplan-Meir curve of survival probability following PEA dependent on post-operative PAWP.

## Discussion

To our knowledge this is the first multicentre in-depth evaluation of the impact of LVDD on morbidity and mortality following PEA. LVDD is well-reported in CTEPH but conventionally purported to be a result of PH and to resolve with surgery [6,12,13]. Our study supports the notion that, far from an isolated phenomenon, LV dysfunction as evidenced by a PAWP > 15mmHg is seen with relative frequency following PEA despite normalisation of pulmonary vascular haemodynamics. Post-operative PAWP elevation was associated with higher mean pulmonary artery pressure, lower cardiac output and reduced functional, patient-reported and mortality outcomes compared to their normal PAWP counterparts.

Abnormal mitral inflow velocities consistent with LVDD are well-recognised in CTEPH and thought to reflect the negative effects of PH on the shared intraventricular septum and left atrial preload [6,7,8,9]. These measures have been shown to normalise with the amelioration of PH by PEA, though studies have been of limited sample size [6, 12, 13]. In our large-scale PEA cohort, however, we demonstrate 14% of individuals to have an elevated PAWP following PEA, corroborating the recent findings of [14]. Our cardiac imaging data supports the premise that PAWP elevation post-PEA, following the resolution of PH, reflects true impairment in LV diastolic function, in line with epidemiological data in relevant age stratified general populations [15]. Although in current guidelines, the presence of a resting PAWP > 15 mmHg confirms definite evidence of Heart Failure Preserved Ejection Fraction (HFpEF), we have stopped short of labelling these patients as having co-existing HFpEF in recognition of the complex interplay of the left and right sides in this patient group [25]. Explicitly though in naming this group LVDD we are putting them on a continuum with HFpEF.

Pre-operatively CTEPH was associated reduced early diastolic LV filling with reductions in E-wave velocities in our representative echocardiography subpopulation with reversal of the E/A ratio in keeping with impaired LV relaxation. PEA lead to significant reductions across all CMR measures of RV dilatation and hypertrophy and an increase in RV function. Overall, there was normalisation in measures of LV compliance, filling and function across the CTEPH cohort with PEA, alongside a 10% increase in indexed left atrial volume.

Post-operatively 14% of our CTEPH cohort had an elevated PAWP, consistent with other reports [14]. Whilst LVEDP was not directly measured in this study, excellent agreement has previously been demonstrated between PAWP and LVEDP readings in CTEPH [14]. PAWP elevation was observed in the context of preserved LV systolic function, whilst abnormalities in LV diastolic function were seen across cardiac imaging modalities. Post-operative PAWP elevation was associated with higher; left atrial area, mitral inflow and tissue doppler velocities, LV mass and relative wall thickness and lower mitral deceleration times on echocardiography. Whilst on CMR, PAWP elevation was associated with reduced indexed end-systolic and end-diastolic volumes, LV stroke volume and left atrial volume compared to their normal PAWP counterparts. LV mass was also disproportionately raised for LV volume.

PEA resulted in significant improvements in RV dilatation, hypertrophy and function in those with a normal post-operative PAWP. LV filling, compliance and stroke volume also increased significantly in this group. In the elevated PAWP group, whilst CMR RV measures improved with surgery, these changes were attenuated compared to their normal PAWP counterparts, even following the removal of confounders (eg atrial arrhythmias). Furthermore, no significant improvements were observed in any CMR LV measure following PEA. The absence of a significant change in left atrial size with PEA in those with an elevated PAWP contradicts prior presumption that left atrial volumes significantly increase following surgery, even in those with pre-existing left heart disease [16].

Overall outcomes following PEA in our study were excellent consistent with previously published data from our own cohort and that of other high-volume centres [2–4, 19]. However, in those with an elevated PAWP following PEA, improvements in mean pulmonary artery pressure, 6-minute walk distance, NYHA functional class and PRO scores were attenuated compared to their counterparts with low post-operative PAWP; despite equivalent reductions in pulmonary vascular resistance with surgery. Furthermore, improvements in both plasma NTproBNP level and cardiac index were not observed post-PEA in those with post-operative PAWP elevation. This, along with an increased requirement for perioperative non-invasive ventilation in this group, fits conceptually with the inability of a stiff LV to cope with the demands of post-operative pulmonary venous return.

The implications of post-operative LVDD extends beyond that of reduced functional gains with surgery. Our large multicentre dataset is in agreement with Gerges et al, who demonstrated a close agreement between PAWP and LVEDP in operable CTEPH [14]. Our work clarifies that LVDD is a risk factor for both functional and mortality outcomes. A PAWP > 15mmHg remained an independent marker of prognostication in multivariate modelling containing relevant demographic, haemodynamic and functional measures of outcome. This has important ramifications when considering surgical risk/benefit ratio and in the planning of post-surgical care.

Of particular interest is that over half of those with elevated PAWPs following PEA, had a pre-operative PAWP ≤ 15mmHg. Whether this population represents an ‘at risk’ group with cardiovascular risk factors who go on to develop LVDD as a result of the haemodynamic insult of major cardiothoracic surgery, or whether this population has pre-existing LVDD masked pre-operatively by LV underfilling, is unclear. The absence of association between intra-operative factors including concomitant cardiac procedures, circulatory arrest duration and hypothermic time is however suggestive towards the latter.

More importantly is whether we are able to identify patients who have LVDD prior to operation. We found a number of cardiac risk factors to associate with the technical development of HFpEF (as per guideline definition of PAWP >15mmHg in absence of systolic dysfunction). These included; obesity, atrial arrhythmias, lower functional status and higher pre-operative PAWP itself. The current absence of validated diagnostic algorithms for LVDD in the presence of PH has previously made screening for these ‘at risk’ patients pre-PEA using imaging modalities challenging. However, our study demonstrates that elevated LV filling pressures post-operatively can be predicted prior to surgery with a high degree of accuracy by CMR-derived left atrial volume alone (sensitivity 67%, specificity 100%) with a threshold value of 63mls, outperforming echocardiographic left atrial area (sensitivity 64%, specificity 68%). Whilst standard practice is to adjust atrial measures for body surface area, this unexpectedly resulted in a loss of association with PAWP elevation in our cohort. As half of our population had a BMI ≥ 30kg/m^2^, indexing may have led to an under-estimation of left atrial indices in this group.

This study has a number of strengths; particularly in describing a whole nation’s surgical cohort, thereby limiting referral and population bias. Over-wedging of PAWP is a theoretical confounder in CTEPH, though our echocardiographic and MR data strongly support this not being a confounder. In post-PEA PAWP the bias should be further limited as proximal disease has been removed.

## Conclusion

LVDD in CTEPH may be attributable to the effects of PH but, in a subset of individuals, intrinsic LVDD can be discerned due the unmasking of LV pathology following PEA, due to increased preload. Whilst these patients still derive benefit from surgery compared to their non-operated counterparts, there is clear evidence that LVDD has a negative impact on survival, haemodynamic, functional and PRO gains following PEA. Consideration should be given to the optimisation of patients background cardiovascular treatments and work now needs to focus on clarifying if this impacts on outcomes.

## Data Availability

All data produced in the present work are contained in the manuscript

## Abbreviations

CMR: Cardiac magnetic resonance
CTEPH: Chronic thromboembolic pulmonary hypertension
HFpEF: Heart failure preserved ejection fraction
LV: Left ventricle
LVDD: Left ventricular diastolic dysfunction
LVEDP: Left ventricular end-diastolic pressure
PAWP: Pulmonary arterial wedge pressure
PEA: Pulmonary endarterectomy
PH: Pulmonary hypertension
PRO: Patient Reported Outcome
PVR: Pulmonary vascular resistance
RV: Right ventricle

## Author contributions

K.B and M.N collected and curated the clinical data. KB performed the analysis, interpreted data and wrote the manuscript. M.T supervised the study. A.B, R.V.G and J.W-M analysed and curated the cardiac MR data. A.F contributed to echocardiogram data collection and curation. G.C, J.L, L.H, D.J. M.J, D.K, C.N. K.S, D.T, S.T, S.J.W, J.P-Z contributed to data collection. All authors contributed significantly to the writing and review of the main manuscript, had full access to all the data in the study, and had final responsibility for the decision to submit the manuscript for publication.

