## Supplemental data for "Left ventricular diastolic dysfunction attenuates outcomes in chronic thromboembolism pulmonary hypertension"

Online data supplement

**Supplementary Tables**

**Table S1**: Cohort echocardiographic variables pre- and post-PEA

|  | n | **Pre-PEA** | n | **Post-PEA** | **p** |
| --- | --- | --- | --- | --- | --- |
| Left atrial area, cm^2^ | 263 | 16.6 ± 4.0 | 233 | 19.0 ± 5.9 | < 0.001 |
| Right atrial area, cm^2^ | 255 | 24.2 ± 11.3 | 242 | 20.2 ± 8.2 | < 0.001 |
| RV basal diameter, cm | 220 | 4.7 ± 1.1 | 216 | 4.1 ± 1.1 | < 0.001 |
| TR velocity, m/s | 108 | 388 ± 84 | 102 | 265 ± 83 | < 0.001 |
| TAPSE, mm | 268 | 18 ± 7 | 266 | 15 ± 1 | < 0.001 |
| Systolic PAP, mmHg | 251 | 77 ± 28 | 231 | 39 ± 16 | < 0.001 |
| PAT, msec | 115 | 77 ± 22 | 159 | 100 ± 29 | < 0.001 |
| Systolic LV EI | 235 | 1.4 ± 0.5 | 232 | 1.0 ± 0.1 | < 0.001 |
| Diastolic LV EI | 237 | 1.3 ± 0.3 | 214 | 1.0 ± 0.0 | < 0.001 |
| LVEF, % | 262 | 64 ± 5 | 279 | 60 ± 6 | 0.004 |
| LV mass index, g/m^2^ | 170 | 77.2 ± 26.2 | 156 | 73.8 ± 23.5 | 0.579 |
| LV RWT | 136 | 0.43 ± 0.12 | 128 | 0.41 ± 0.11 | 0.083 |
| Mitral E, cm/s | 260 | 55.6 ± 25.1 | 302 | 65.7 ± 32.5 | < 0.001 |
| Mitral A, cm/s | 240 | 70.0 ± 22.9 | 210 | 72.1 ± 28.9 | < 0.001 |
| Mitral E/A ratio | 240 | 0.75 ± 0.29 | 210 | 0.85 ± 0.47 | < 0.001 |
| Mitral lateral e´, cm/s | 187 | 10.4 ± 4.2 | 208 | 10.8 ± 4.7 | 0.457 |
| Mitral lateral E/e´ | 183 | 5.1 ± 2.3 | 210 | 6.6 ± 3.2 | < 0.001 |
| Mitral average E/e´ | 197 | 7.3 ± 2.9 | 171 | 8.0 ± 3.4 | 0.005 |
| Mitral DT, ms | 121 | 230 ± 98 | 202 | 206 ± 77 | 0.003 |

Values expressed as median ± IQR. Pre-PEA values taken at time of diagnostic right heart catheterisation. Post-PEA values taken at first follow-up within one year of PEA.

PEA, pulmonary endarterectomy; RV, right ventricle; TR, tricuspid regurgitation; TAPSE, tricuspid annular plane excursion; PAP, pulmonary artery pressure; PAT, pulmonary acceleration time; LV, left ventricle; EI, eccentricity index; LVEF, left ventricular ejection fraction; RWT, relative wall thickness; DT, deceleration time.

**Table S2**: Cohort haemodynamic, functional and self-reported measures pre- and post- PEA (unpaired).

|  | n | **Pre-PEA** | n | **Post-PEA** | P |
| --- | --- | --- | --- | --- | --- |
| **Total n** | 1266 |  | 1177 |  |  |
| **Haemodynamics** |  |  |  |  |  |
| Mean PAP, mmHg | 1266 | 45 ± 15 | 1022 | 25 ± 13 | < 0.001 |
| PVR, dynes | 1202 | 676 ± 485 | 997 | 246 ± 218 | < 0.001 |
| CI, l/min/m2 | 1125 | 2.1 ± 0.75 | 985 | 2.3 ± 0.72 | < 0.001 |
| **Functional status** |  |  |  |  |  |
| NYHA class, 1/2/3/4% | 1088 | 0/15/74/11 | 944 | 29/43/26/1 | < 0.001 |
| 6MWD, metres | 732 | 316 ± 195 | 944 | 365 ± 162 | < 0.001 |
| **CAMPHOR** |  |  |  |  |  |
| Symptoms | 1187 | 12 ± 11 | 944 | 4 ± 9 | < 0.001 |
| Activity | 1187 | 11 ± 10 | 944 | 6 ± 10 | < 0.001 |
| Quality of Life | 1187 | 10 ± 12 | 944 | 4 ± 11 | < 0.001 |

Values are expressed as median ± IQR. % may not add to 100 due to rounding. Pre-PEA values taken at diagnostic right heart catheterisation. Post-PEA outcomes taken from first follow-up within one year of PEA. P-values are corrected for multiple comparison by false discovery rate at 5%.

PEA, Pulmonary Endarterectomy; PAP, pulmonary artery pressure; PVR, pulmonary vascular resistance; CI, cardiac index; NYHA, New York Heart Association functional class; 6MWD, 6-minute walk distance; CAMPHOR, Cambridge Pulmonary Hypertension Outcome Review.

**Table S3:** Comparison of patient characteristics and variables for cohort and echocardiographic subpopulation

|  | n | **Cohort** | n | **Echocardiography subpopulation** |
| --- | --- | --- | --- | --- |
| **Total n** |  | 1266 |  | 311 |
| **Age at PEA**, yrs | 1266 | 62 ± 22 | 311 | 62 ± 20 |
| **Sex,** Male (%) | 1266 | 54 | 311 | 57 |
| **BMI**, kg/m^2^ | 909 | 29 ± 8 | 277 | 28 ± 8 |
| **Comorbidities** |  |  |  |  |
| Atrial arrhythmia, % | 1227 | 9 | 308 | 11 |
| Systemic hypertension, % | 1227 | 27 | 308 | 26 |
| Type 2 Diabetes Mellitus, % | 1227 | 11 | 308 | 13 |
| IHD^b^, % | 1227 | 12 | 311 | 11 |
| **Haemodynamics** |  |  |  |  |
| Mean PAP, mmHg | 1266 | 45 ± 15 | 311 | 44 ± 16 |
| PAWP, mmHg | 1202 | 11 ± 5 | 303 | 11 ± 5 |
| PVR, dynes | 1052 | 676 ± 485 | 306 | 707 ± 479 |
| CI, l/min/m2 | 1125 | 2.1 ± 0.8 | 290 | 1.9 ± 0.6 |
| **Functional status** |  |  |  |  |
| NYHA, 1/2/3/4% | 1088 | 0/15/74/11 | 282 | 0/18/71/11 |
| 6MWD, metres | 732 | 316 ± 195 | 274 | 316 ± 169 |
| **Additional procedures**, % | 1266 | 13 | 311 | 11 |
| **CPB time**, mins | 1266 | 321 ± 65 | 298 | 321 ± 70 |
| **DHCA time**, mins | 1266 | 37 ± 16 | 298 | 37 ± 16 |

Values are expressed as median ± IQR. % may not add to 100 due to rounding. Variables taken at time of diagnostic right heart catheterisation.

PEA, pulmonary endarterectomy; BMI, body mass index; IHD, ischaemic heart disease; PAP, pulmonary artery pressure; PAWP, pulmonary arterial wedge pressure; PVR, pulmonary vascular resistance; CI, cardiac index; NYHA, New York Heart Association functional class; 6MWD, 6-minute walk distance; CPB, cardiopulmonary bypass; DHCA, deep hypothermic circulatory arrest.

^a^ At any time point prior to PEA ^b^History of angina, coronary artery stenting, use of nitrates or coronary artery bypass grafting required at time of pulmonary endarterectomy

**Table S4:** Comparison of patient characteristics and variables for cohort and CMR subpopulation

|  | n | **Cohort** | n | **CMR subpopulation** |
| --- | --- | --- | --- | --- |
| **Total n** |  | 1266 |  | 142 |
| **Age at PEA**, yrs | 1266 | 62 ± 22 | 142 | 63 ± 19 |
| **Sex,** Male (%) | 1266 | 54 | 142 | 63 |
| **Comorbidities** |  |  |  |  |
| Atrial arrhythmia^a^, % | 1227 | 9 | 139 | 11 |
| Systemic hypertension, % | 1227 | 27 | 139 | 26 |
| Type 2 Diabetes Mellitus, % | 1227 | 11 | 139 | 8 |
| IHD^b^, % | 1227 | 12 | 142 | 8 |
| **Haemodynamics** |  |  |  |  |
| Mean PAP, mmHg | 1266 | 45 ± 15 | 142 | 45 ± 16 |
| PAWP, mmHg | 1202 | 11 ± 5 | 130 | 11 ± 6 |
| PVR, dynes | 1052 | 676 ± 485 | 136 | 731 ± 567 |
| CI, l/min/m^2^ | 1125 | 2.1 ± 0.8 | 135 | 1.9 ± 0.7 |
| **Functional status** |  |  |  |  |
| NYHA, 1/2/3/4 % | 1088 | 0/15/74/11 | 131 | 0/21/71/8 |
| 6MWD, metres | 732 | 316 ± 195 | 119 | 308 ± 152 |
| **Additional procedures**, % | 1266 | 13 | 142 | 8 |
| **CPB time**, mins | 1266 | 321 ± 65 | 142 | 334 ± 65 |
| **DHCA time**, mins | 1266 | 37 ± 16 | 142 | 37 ± 17 |

**Table S5:** Cohort CMR variables pre- and post-PEA

|  | **Pre-PEA**  **(n = 142)** | **Post-PEA**  **(n = 142)** | **p-value** |
| --- | --- | --- | --- |
| **Right atrium** |  |  |  |
| Volume index, mls | 52 ± 49 | 38 ± 23 | < 0.001 |
| **Right ventricle** |  |  |  |
| End-diastolic volume index, mls/m^2^ | 108 ± 48 | 75 ± 23 | < 0.001 |
| End-systolic volume index, mls/m^2^ | 70 ± 45 | 79 ± 16 | < 0.001 |
| Ejection fraction, % | 36 ± 18 | 46 ± 14 | < 0.001 |
| Stroke volume index, mls/m^2^ | 37 ± 14 | 35 ± 14 | 0.046 |
| Mass index, g/m^2^ | 27 ± 11 | 20 ± 5 | < 0.001 |
| **Left atrium** |  |  |  |
| Volume index, mls/m^2^ | 29 ± 16 | 32 ± 16 | < 0.001 |
| Ejection fraction, % | 60 ± 15 | 55 ± 13 | 0.002 |
| **Left ventricle** |  |  |  |
| End-diastolic volume index, mls/m^2^ | 65 ± 45 | 73 ± 16 | < 0.001 |
| End-systolic volume index, mls/m^2^ | 29 ± 14 | 31 ± 11 | 0.032 |
| Ejection fraction, % | 57 ± 11 | 55 ± 10 | 0.041 |
| Cardiac output, l/min | 5.6 ± 1.7 | 5.8 ± 1.7 | < 0.001 |
| Stroke volume index, mls/m^2^ | 36 ± 14 | 39 ± 12 | 0.032 |
| Mass index, g/m^2^ | 52 ± 11 | 55 ± 17 | 0.017 |
| Mass/volume g/mls | 0.80 ± 0.22 | 0.76 ± 0.19 | 0.009 |

**Table E6**: Patient characteristics of patients with elevated PAWP post-PEA stratified by pre-operative PAWP

|  | n | **Pre-PEA**  **PAWP** ≤ **15mmHg** | n | **Pre-PEA**  **PAWP > 15mmHg** | p |
| --- | --- | --- | --- | --- | --- |
| **Total n** |  | 42 |  | 63 |  |
| Age, yrs | 42 | 60 ± 20.5 | 63 | 64 ± 20.5 | 0.538 |
| Male sex, n (%) | 42 | 20 (48) | 63 | 38 (60) | 0.538 |
| BMI, kg/m2 | 42 | 32 ± 6 | 63 | 35 ± 14 | 0.171 |
| **Haemodynamics** |  |  |  |  |  |
| Mean PAP, mmHg | 42 | 50 ± 12.5 | 63 | 44 ± 15.5 | 0.171 |
| PVR, dynes | 41 | 485 ± 295 | 63 | 570 ± 421 | 0.286 |
| CI, l/min/m2 | 41 | 2.2 ± 0.6 | 63 | 2.2 ± 0.9 | 1.00 |
| **Functional status** |  |  |  |  |  |
| NYHA class, 1/2/3/4% | 40 | 0/8/80/13 | 55 | 0/13/75/13 | 0.967 |
| 6MWD, metres | 17 | 246 ± 176 | 42 | 245 ± 211 | 0.967 |
| **CAMPHOR** |  |  |  |  |  |
| Activity | 39 | 11 ± 12 | 61 | 15 ± 11 | 0.514 |
| Symptoms | 39 | 12 ± 13 | 61 | 15 ± 12 | 0.514 |
| Quality of Life | 39 | 8 ± 12 | 61 | 12 ± 9 | 0.542 |
| **Co-morbidities** |  |  |  |  |  |
| Atrial arrhythmia, n (%) | 41 | 12 (29) | 63 | 6 (10) | 0.966 |
| Systemic hypertension, n (%) | 41 | 10 (24) | 63 | 6 (10) | 1.00 |
| IHD, n (%) | 41 | 20 (49) | 63 | 13 (21) | 1.00 |
| Type 2 DM, n (%) | 41 | 9 (22) | 63 | 13 (21) | 0.287 |
| **Intra-operative variables** |  |  |  |  |  |
| Additional procedures, n (%) | 42 | 8 (19) | 63 | 11 (17) | 1.00 |
| CPB time, mins | 34 | 332 ± 60 | 61 | 326 ± 67 | 0.966 |
| DHCA time, mins | 40 | 35 ± 13 | 62 | 13 ± 10 | 0.287 |

Values are expressed as median ± IQR. % may not add to 100 due to rounding. Post-PEA outcomes taken from first follow-up within one year of PEA. PEA, Pulmonary Endarterectomy; BMI, Body Mass Index; PAP, pulmonary artery pressure; PVR, pulmonary vascular resistance; CI, cardiac index; NYHA, New York Heart Association functional class; 6MWD, 6-minute walk distance; CAMPHOR, Cambridge Pulmonary Hypertension Outcome Review; IHD, ischaemic heart disease; DM, diabetes mellitus; CPB, cardiopulmonary bypass; DHCA, deep hypothermic circulatory arrest.

**Table S7**: Echocardiographic variables at first post-operative RHC stratified by left atrial pressure (individuals with underlying atrial arrhythmias removed)

|  | n | **PAWP** ≤ **15mmHg** | n | **PAWP > 15mmHg** | **p** |
| --- | --- | --- | --- | --- | --- |
| Total n |  | 233 |  | 23 |  |
| Left atrial area, cm^2^ |  | 18.0 ± 5.1 |  | 22.6 ± 1.9 | 0.017 |
| Right atrial area, cm^2^ |  | 19.1 ± 7.9 |  | 23.1 ± 7.2 | 0.049 |
| TR velocity, m/s |  | 261 ± 79 |  | 310 ± 38 | 0.053 |
| LVEF, % |  | 60 ± 6.0 |  | 60 ± 9.5 | 0.784 |
| LV mass index, g/m^2^ |  | 73.3 ± 24.3 |  | 88.6 ± 15.8 | 0.049 |
| LV RWT |  | 0.40 ± 0.11 |  | 0.50 ± 0.08 | 0.049 |
| Mitral E, cm/s |  | 62.8 ± 28.2 |  | 87.2 ± 34.5 | 0.017 |
| Mitral A, cm/s |  | 70.4 ± 27.0 |  | 97.0 ± 28.4 | 0.049 |
| Mitral E/A ratio |  | 0.84 ± 0.48 |  | 0.89 ± 0.43 | 0.427 |
| Mitral lateral e´, cm/s |  | 10.9 ± 4.7 |  | 10.0 ± 3.9 | 0.728 |
| Mitral lateral E/e´ |  | 6.0 ± 3.1 |  | 7.8 ± 2.1 | 0.017 |
| Mitral average E/e´ |  | 8.0 ± 3.1 |  | 10.0 ± 4.2 | 0.049 |
| Mitral DT, ms |  | 210 ± 77 |  | 170 ± 50 | 0.032 |

Values expressed as median ± IQR. P-values are adjusted for multiple comparisons at FDR 5%.

PAWP, pulmonary artery wedge pressure; TR, triscuspid regurgitation; LVEF, Left ventricular ejection fraction; LV, left ventricle; RWT, relative wall thickness; DT, deceleration time.

**Table S8:** CMR variables for individuals with a post-operative PAWP ≤ 15mmHg (n = 119)

|  | **Pre PEA** | **Post PEA** | **p-value** |
| --- | --- | --- | --- |
| **Right atrium** |  |  |  |
| Volume index, mls/m^2^ | 52 ± 50 | 38 ± 24 | < 0.001 |
| **Right ventricle** |  |  |  |
| End-diastolic volume index, mls/m^2^ | 107 ± 43 | 77 ± 22 | < 0.001 |
| End-systolic volume index, mls/m^2^ | 70 ± 40 | 40 ± 15 | < 0.001 |
| Ejection fraction, % | 36 ± 19 | 47 ± 13 | < 0.001 |
| Cardiac output, l/min | 5.7 ± 2.1 | 5.4 ± 1.9 | < 0.001 |
| Stroke volume index, mls/m^2^ | 38 ± 13 | 38 ± 14 | 0.843 |
| Mass index, g/m^2^ | 27 ± 10 | 20 ± 5 | < 0.001 |
| **Left atrium** |  |  |  |
| Volume index, mls/m^2^ | 28 ± 19 | 32 ± 18 | < 0.001 |
| Ejection fraction, % | 60 ± 15 | 57 ± 10 | 0.005 |
| **Left ventricle** |  |  |  |
| End-diastolic volume index, mls/m2 | 67 ± 20 | 76 ± 17 | < 0.001 |
| End-systolic volume index, mls/m^2^ | 29 ± 8 | 33 ± 13 | < 0.001 |
| Ejection fraction, % | 56 ± 10 | 55 ± 10 | 0.141 |
| Cardiac output, l/min | 5.8 ± 2.1 | 5.9 ± 1.7 | < 0.001 |
| Stroke volume index, mls/m^2^ | 37 ± 15 | 41 ± 12 | 0.018 |
| Mass index, g/m^2^ | 52 ± 12 | 55 ± 18 | 0.046 |
| Mass/volume, g/mls | 0.79 ± 0.22 | 0.75 ± 0.19 | 0.003 |

**Table S9:** CMR variables for individuals with a post-operative PAWP > 15mmHg (n = 14)

|  | **Pre PEA** | **Post PEA** | **p-value** |
| --- | --- | --- | --- |
| **Right atrium** |  |  |  |
| Volume index, mls/m^2^ | 55 ± 29 | 44 ± 25 | 0.544 |
| **Right ventricle** |  |  |  |
| End-diastolic volume index, mls/m2 | 103 ± 63 | 86 ± 22 | 0.165 |
| End-systolic volume index, mls/m^2^ | 58 ± 65 | 50 ± 25 | 0.120 |
| Ejection fraction, % | 38 ± 13 | 38 ± 9 | 0.937 |
| Cardiac output, l/min | 5.8 ± 2.1 | 6.0 ± 2.5 | 0.625 |
| Stroke volume index, mls/m^2^ | 38 ± 13 | 35 ± 13 | 0.600 |
| Mass index, g/m^2^ | 28 ± 7 | 21 ± 3 | 0.120 |
| **Left atrium** |  |  |  |
| Volume index, mls/m^2^ | 37 ± 13 | 36 ± 11 | 0.733 |
| Ejection fraction, % | 56 ± 18 | 45 ± 8 | 0.733 |
| **Left ventricle** |  |  |  |
| End-diastolic volume index, mls/m2 | 69 ± 15 | 59 ± 9 | 0.600 |
| End-systolic volume index, mls/m^2^ | 30 ± 9 | 29 ± 10 | 0.866 |
| Ejection fraction, % | 58 ± 11 | 56 ± 8 | 0.625 |
| Cardiac output, l/min | 5.4 ± 1.4 | 5.5 ± 2.1 | 0.711 |
| Stroke volume index, mls/m^2^ | 36 ± 3 | 34 ± 6 | 0.600 |
| Mass index, g/m^2^ | 58 ± 7 | 60 ± 12 | 0.600 |
| Mass/volume, g/mls | 0.79 ± 0.19 | 0.94 ± 0.42 | 0.499 |

**Table S10:** Change in variable following PEA stratified by post-operative PAWP

|  | **PAWP** ≤ **15mmHg** | | **PAWP > 15mmHg** | |
| --- | --- | --- | --- | --- |
| **Total n** | 862 | | 135 | |
| **Haemodynamics** |  |  |  |  |
| Mean PAP, mmHg | -18 ± 17 | (p < 0.001) | -9 ± 14 | (p < 0.001) |
| PVR, dynes | -362 ± 443 | (p < 0.001) | -228 ± 379 | (p < 0.001) |
| CI, l/min/m2 | 0.21 ± 0.88 | (p < 0.001) | -0.03 ± 0.78 | (p = 0.393) |
| **Functional status** |  |  |  |  |
| NYHA, 1/2/3/4% | -1 ± 2 | (p < 0.001) | -1 ± 1 | (p = 0.005) |
| 6MWD, metres | 54 ± 117 | (p < 0.001) | 42 ± 123 | (p = 0.037) |
| **CAMPHOR** |  |  |  |  |
| Symptoms | -6 ± 9 | (p < 0.001) | -3 ± 7 | (p < 0.001) |
| Activity | -3 ± 7 | (p < 0.001) | -1 ± 6 | (p = 0.023) |
| Quality of Life | -3 ± 8 | (p < 0.001) | -1 ± 5 | (p = 0.005) |
| **NTproBNP**, pg/ml | -273 ± 1199 | (p < 0.001) | 136 ± 956 | (p = 0.993) |

Post-PEA outcomes taken from first follow-up within one year of PEA. Values expressed as median ± IQR. P-values are corrected for multiple comparison by false discovery rate at 5%.

PEA, Pulmonary Endarterectomy; PAP, pulmonary artery pressure; PVR, pulmonary vascular resistance; CI, cardiac index; NYHA, New York Heart Association functional class; 6MWD, 6-minute walk distance; CAMPHOR, Cambridge Pulmonary Hypertension Outcome Review; NTproBNP, N-terminal pro-Brain Natriuretic Peptide.

**Table S11:** Comparison of post-PEA measures for those with and without PAWP elevation following PEA

|  | n | **PAWP**  ≤ **15mmHg** | n | **PAWP**  **> 15mmHg** | p |
| --- | --- | --- | --- | --- | --- |
| **Total n** |  | 862 |  | 135 |  |
| **Haemodynamics** |  |  |  |  |  |
| Mean PAP, mmHg | 857 | 23 ± 11 | 133 | 34 ± 10 | < 0.001 |
| PVR, dynes | 863 | 243 ± 211 | 131 | 260 ± 215 | 0.085 |
| CI, l/min/m2 | 815 | 2.3 ± 0.7 | 131 | 2.1 ± 0.6 | < 0.001 |
| **Functional status** |  |  |  |  |  |
| NYHA class, 1/2/3/4% | 748 | 33/43/23/1 | 114 | 13/42/42/1 | < 0.001 |
| 6MWD, metres | 774 | 376 ± 155 | 112 | 289 ± 174 | < 0.001 |
| **CAMPHOR** |  |  |  |  |  |
| Symptoms | 755 | 4 ± 8 | 117 | 7 ± 10 | < 0.001 |
| Activity | 755 | 6 ± 9 | 117 | 9 ± 12 | < 0.001 |
| Quality of Life | 755 | 3 ± 10 | 117 | 7 ± 10 | < 0.001 |
| **NTproBNP levels**, pg/ml | 331 | 179 ± 320 | 44 | 570 ± 1254 | < 0.001 |

Values are expressed as median ± IQR. % may not add to 100 due to rounding. Post-PEA outcomes taken from first follow-up within one year of PEA.

PEA, Pulmonary Endarterectomy; PAP, pulmonary artery pressure; PVR, pulmonary vascular resistance; CI, cardiac index; NYHA, New York Heart Association functional class; 6MWD, 6-minute walk distance; CAMPHOR, Cambridge Pulmonary Hypertension Outcome Review; NTproBNP, N-terminal pro-Brain Natriuretic Peptide.

**Table S12**: Patient characteristics and variables for survivors and non-survivors to first post-PEA follow-up

|  | n | **Survivors** | n | **Non-survivors** | **P** |
| --- | --- | --- | --- | --- | --- |
| **Total n** |  | 1266 |  | 89 |  |
| **Age at PEA**, yrs | 1266 | 62 ± 22 | 89 | 70 ± 19 | < 0.001 |
| **Sex,** Male (%) | 1266 | 54 | 89 | 44 | 0.103 |
| **Haemodynamics** |  |  |  |  |  |
| Mean PAP, mmHg | 1266 | 45 ± 15 | 89 | 46 ± 13 | 0.638 |
| PAWP, mmHg | 1052 | 11 ± 5 | 74 | 12 ± 5 | 0.456 |
| PVR, dynes | 1202 | 676 ± 485 | 84 | 730 ± 495 | 0.344 |
| CI, l/min/m2 | 1125 | 2.1 ± 0.75 | 86 | 2.1 ± 0.9 | 0.230 |
| **Functional status** |  |  |  |  |  |
| NYHA, 1/2/3/4% | 1088 | 0/15/74/11 | 84 | 0/2/81/17 | 0.028 |
| 6MWD, metres | 732 | 316 ± 195 | 50 | 172 ± 191 | < 0.001 |
| **CAMPHOR** |  |  |  |  |  |
| Symptoms | 1187 | 12 ± 11 | 81 | 15 ± 12 | 0.104 |
| Activity | 1187 | 11 ± 10 | 81 | 15 ± 12 | 0.002 |
| Quality of Life | 1187 | 10 ± 12 | 81 | 14 ± 13 | 0.011 |
| **Additional procedures**, % | 1266 | 13 | 89 | 20 | 0.103 |
| **CPB time**, mins | 1266 | 321 ± 65 | 89 | 332 ± 76 | 0.104 |
| **DHCA time**, mins | 1266 | 37 ± 16 | 89 | 41 ± 24 | 0.103 |

Values are expressed as median ± IQR. % may not add to 100 due to rounding. Post-PEA outcomes taken from first follow-up within one year of PEA. P-values are corrected for multiple comparison by false discovery rate at 5%.

^a^ Additional procedures = concomitant coronary artery bypass grafting, mitral valve replacement, aortic valve replacement or atrial septal defect/patent foramen ovale repair at the time of PEA

PEA, Pulmonary Endarterectomy; PAP, pulmonary artery pressure; PAWP, pulmonary arterial wedge pressure; PVR, pulmonary vascular resistance; CI, cardiac index; NYHA, New York Heart Association functional class; 6MWD, 6-minute walk distance; CAMPHOR, Cambridge Pulmonary Hypertension Outcome Review; CPB, cardiopulmonary bypass; DHCA, deep hypothermic circulatory arrest.

**Table S13**: Cox Proportional Hazards model for post-PEA survival (n = 1177)

|  | | **Univariate** | | **Multivariate** | |
| --- | --- | --- | --- | --- | --- |
|  | HR [95% CI] | | p-value | HR [95% CI] | p-value |
| Year of PEA | 1.01 [0.93 – 1.09] | | 0.791 | - | - |
| Age | 1.05 [1.03 – 1.07] | | < 0.001 | 1.04 [1.02 – 1.06] | 0.001 |
| Sex | 1.35 [0.90 – 2.02] | | 0.143 | - | - |
| Mean PAP | 1.05 [1.03 – 1.07] | | < 0.001 | ^a^ | ^a^ |
| PVR | 1.00 [1.00 – 1.00] | | < 0.001 | 1.04 [0.99 – 1.09] | 0.082 |
| Cardiac Index | 0.41 [0.26 – 0.65] | | < 0.001 | 0.27 [0.45 – 1.58] | 0.146 |
| PAWP > 15mmHg | 1.86 [1.07 – 3.25] | | < 0.001 | 2.01 [1.16 – 3.48] | 0.012 |
| 6-MWD | 0.99 [0.99 – 1.00] | | < 0.001 | 0.41 [0.28 – 0.59] | 0.029 |

PEA, pulmonary endarterectomy; PAP, pulmonary artery pressure; PVR, pulmonary vascular resistance; PAWP, pulmonary arterial wedge pressure; 6MWD, six-minute walk distance.

^a^ mean PAP removed from multivariate analysis due to collinearity with PAWP (r = 0.49, p < 0.001)

**Figures**

**Figure S1:** Paired PAWP measurements pre- and post-PEA (n = 836**)**

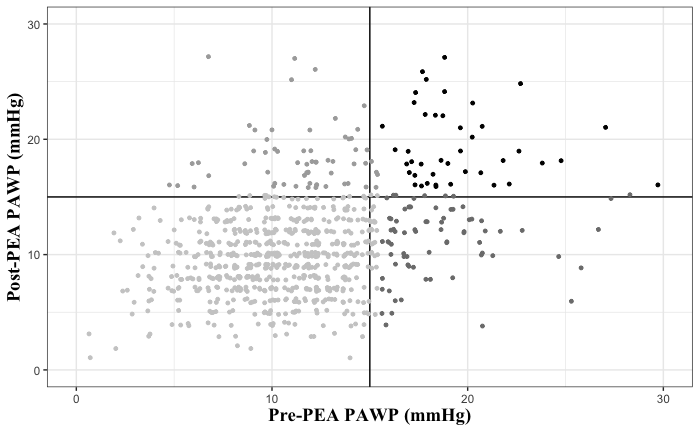

n = 653

n = 78

n = 42

n = 63

Pre-PEA PAWP taken at diagnostic right heart catheterisation. Post-PEA PAWP taken at first follow-up right heart catheterisation within one year of PEA.

PEA, pulmonary endarterectomy

**Cardiac imaging additional methodology**

Echocardiographic images were obtained using Philips (Amsterdam, Netherlands) IE33 and Epiq ultrasound machines and analysed with Philips Xcelera/Intelispace software

Parameters derived from CMR were analysed for a subset of the cohort comprising all individuals with available paired pre- and post-operative CMR data (n = 142). Images were stored on the local imaging system Cadran Picture Archiving System (PACS). CMR images were obtained at the referring centres using 1.5-T cardiac optimised system (multivendor; Siemens Medical Solutions, Erlangen, Germany). Volumetric analysis was calculated from balanced Steady State Free Precession (bSSFP) technique cine imaging. A three-plane localiser and axial half-Fourier acquisition turbo spin echo (HASTE), were used to derive the standard cardiac planes: long axis 2-chamber, 3-chamber and 4-chamber views and a short axis ventricular stack. ECG-triggering was used to obtain over the whole cardiac cycle. A line was drawn through the atrioventricular groove in the 4-chamber and 2-chamber long axis cine sequence to produce a bSSFP stack of short axis cine-images starting from the most basal down to the most apical section of the ventricles (the following values are representative but may vary slightly between referring centres: slice thickness= 8mm, interslice gap= 2mm, trigger time (TD) 260.70, repetition time (TR) 43.65, echo time (TE) 1.24, inversion time (TI), phases = 40, pixel spacing 1.25, 1.25, , flip angle 75, percent phase field of view (FOV) 92.5). Breath-holding was used to reduce motion artefact. ECG-triggering was used to obtain images in end-diastole and end-systole segment the images to cover the whole cardiac cycle. Image post-processing was performed using a Machine Learning software using a tensor-based approach with multilinear subspace learning previously trained in a multicentre multivendor external cohort and validated on the study population [Alandejani et al., 2022]. This defined endocardial and epicardial contours throughout the entirety of the cardiac cycle on both long- and short-axis views. All cines were reviewed to ensure accurate contouring throughout the cardiac cycle and adjusted where necessary. Ventricular volumes were measured calculated from the short axis stack, while 2-chamber and 4-chamber view and atrial volumes were calculated from the 4-chamber views. Right ventricular endocardial borders were drawn on the middle of the chemical shift artefact. The trabeculae and papillary muscles were included within the borders of the ventricular cavity volumes. The left and right ventricular outflow tracts were included with the endocardial contour drawn up to the level of aortic and pulmonary valve cusps. The end-diastole was defined as the cardiac slice phase with the largest volume and end-systole as the slide phase with the lowest volume (calculated separately for the left and right ventricular volumes to account for inter-ventricular dysynchrony). From these, the RV and LV end-diastolic volume (EDV), end-systolic volume (ESV), LV stroke-volume (SV), and ejection fraction (EF) and LV mass were generated. LV Radial and circumferential endocardial and wall strain were derived from 2-chamber short axis cine views at the mid papillary ventricular level. Longitudinal strain was derived from the 4-chamber views.
